# Mobile Phone Virtual Reality Game for Pediatric Home Burn Dressing Pain Management, a Randomized Clinical Trial

**DOI:** 10.1101/2022.01.07.22268893

**Authors:** Megan Armstrong, Jonathan Lun, Jonathan I. Groner, Rajan K. Thakkar, Renata Fabia, Dana Noffsinger, Henry Xiang

**Author notes:** **Corresponding author:** Henry Xiang, MD, MPH, PhD, MBA, Director of Center for Pediatric Trauma Research & Director of Research Core/Center for Injury Research and Policy, Nationwide Children’s Hospital, 700 Children’s Drive, Columbus, Ohio 43205. **Declarations of Interest:** None.

## Abstract

**Importance:** Virtual Reality (VR) gaming is considered a safe and effective alternative to standard pain alleviation in the hospital, we advocate for its use during repeated redressing at home.

**Objective:** This study will address the effectiveness and feasibility of the Virtual Reality Pain Alleviation Tool (VR-PAT) that was developed by the research team at the Nationwide Children’s Hospital for repeated at-home burn dressing changes.

**Design, Setting and Participants:** Randomized clinical trial among patients recruited at the Nationwide Children’s Hospital (NCH) outpatient burn clinic between September 2019 and June 2021. We included English-speaking burn patients 5-17 years old requiring daily dressing changes for at least one week after first outpatient dressing change.

**Interventions:** One group played an interactive VR-PAT game during dressing changes, while the other utilized standard distraction techniques available in the home for a week. Both child and caretaker were later asked to assess perceived pain. Those in the intervention group were asked to evaluate convenience and enjoyment of the VR-PAT game.

**Outcomes:** Patients were asked to rate perceived pain on a scale of 1-10, and caregivers were asked to rate observed pain on a scale of 1-10. For the VR-PAT group, patients were also asked to rate various aspects of the VR game on a scale of 1-10 and caregivers were asked questions assessing ease of use.

**Conclusions:** Subjects found the VR-PAT to be a useful distraction during home dressing changes and reported it be easy to implement. In the VR-PAT group, child and caregiver reported pain decreased as the week of dressing changes progressed and was lower than those in the control group after the fourth dressing change. Children playing the VR-PAT reported consistent happiness and fun as the week went on and increased realism and engagement, which means our results weren’t just due to the novel experience of VR-PAT.

**Trial Registration:** ClinicalTrials.gov Identifier: NCT04548635

## Introduction

Since the turn of the century, treatment options for pediatric burn patients have taken significant strides. This is evidenced by a 48% decrease in mortality and 64% increase in patient referral to larger, more well-equipped institutions in the past 20 years.^1^ However, pediatric burns continue to be a serious public health issue. In 2019, the CDC reported that burns are the fourth leading cause of death due to unintentional injury in children ages 1-14.^2^ A major area for improvement is in the field of pain management. This includes acute/nociceptive pain from both the initial burn and procedural pain from subsequent dressing changes.^3^

Dressing changes are often more painful than the initial burn and may require high-dose opioids.^4^ This is particularly concerning, as studies show a nationwide two-fold increase in pediatric mortality rate due to opioid poisonings since the early 2000’s.^5^ The danger of opioids must be balanced with the need for adequate pain relief. Repeated dressing-associated pain has been shown to increase anxiety and learned helplessness in pediatric patients.^6,7^ As a result, nonpharmacologic alternatives have risen to the forefront of pain management research. Methods such as hypnosis, cognitive behavioral therapy and distraction are now standard protocol in the hospital setting.^8^

Virtual Reality (VR) provides a much more immersive distraction than standard techniques such as muscle relaxation and toys.^9^ Multiple studies have demonstrated that VR distraction is clinically beneficial when compared with current standard of care.^9-11^ In addition to their efficacy, VR games are also rated by patients as enjoyable, user friendly and having few side effects.^9^ VR as a pain alleviation tool for burn victims is well-studied in the hospital, however there is little literature focusing on its use during at-home dressing changes. Furthermore, previous studies investigating VR gaming as a pain management dated back to 1980 and many of them used the bulky computer-based systems, which is not very practical for clinical implementation as well as at-home burn dressing change.

The lack of studies pertaining to the use of VR during at-home burn management is surprising, as recent advances in technology have allowed VR to be accessible, relatively inexpensive and common in gaming, education and medical care.^12^ From a clinical perspective, at-home VR has demonstrated utility in treatment of public speaking anxiety^13^, fall prevention in the elderly^14^ and neuromuscular rehabilitation.^15^ VR has also shown promise in terms of managing chronic pain. A 2017 study by Garrett et al. found that at-home VR therapy was effective in reducing short-term pain among patients with chronic conditions.^16^ The diverse use of VR in at-home care combined with its promise as a nonpharmacologic pain management tool makes it an ideal potential adjunct to standard home burn care.

National statistics reported that over 250,000 US children (0-17 years) suffer burn injuries every year.^17^ Over half of pediatric burn injuries seen in US emergency departments (EDs) are serious enough to merit referral to a burn center according to US and international guidelines.^8,18^ After being discharged from medical burn care facilities, at-home repeated burn dressing changes are often needed for 2-3 weeks. Dressing changes have been identified by pediatric patients as very painful, with opioid and anxiety medications often being prescribed.^19,20^ Furthermore, the pain experienced during burn dressing changes may cause distress to not only pediatric patients but also their caregivers.^21^ A painful experience can additionally serve as a stressor that significantly impacts patients’ post-injury health outcomes.^22,23^ The latest research shows that repeated use of opioid medication for acute pain management is likely to increase the risk of long-term opioid use and risk of opioid addictions.^24,25^ The medical community in the US is diligently working to find the right balance between the risk of undertreating pain and causing unneeded suffering^19,20^ and the risk of over (or inappropriate) prescription of opioids.^26^ The “50 State Review on Opioid Related Policy”, published in late 2017, reported that 11 states have laws or guidelines in place that encourage the use of non-opioid alternatives for pain treatment.^27^ The Ohio State Medical Board mandates that all prescriptions will need to carry documentation which “should indicate whether there are known and available non-opiate alternatives and why it has been determined not to utilize these alternatives”. Therefore, there is a pressing need to seek non-pharmacological interventions for effective pain management in pediatric burn wound care.

Non-pharmacological pain management is widely utilized by pediatric clinicians as safe and affordable solutions for procedural acute pain management.^28,29^ According to the Cognitive-Affective Model of Pain,^30^ pain perception demands cognitive attention. Thus, effective non-pharmacological pain management requires the interruption of the cognitive route from the origin to pain perception, redirecting (distracting) a child’s attention resources away from the painful procedure. VR has been demonstrated to effectively decrease pain and distress in a variety of settings, diverse populations (i.e. children as well as adults),^31,32^ and a wide range of pain conditions (i.e. acute, procedural, and chronic).^33^ Researchers have provided preliminary but convincing evidence that suggests the neurobiological interplay of brain cortices and neurochemistry, as well as emotional, cognitive, and attentional processes as the underlying mechanistic origin for VR analgesia.^34^ Gold et al. and other researchers have demonstrated that VR reduces pain signaling in this pain matrix that mimics, in part, the actions of opiates.^33-36^

Prior research confirmed that VR provides three unique advantages over traditional non-pharmacological interventions for pediatric burn patients.^9,37^ First, VR technology is capable of creating a three-dimensional immersive virtual environment (e.g. visual; auditory; interaction) for actively engaging the pediatric patients’ attention in order to successfully interrupt the pain perception route, consistent with the Cognitive-Affective Model of Pain^29,38^ and underlying mechanisms of pain management. The unique highly immersive experience of presence, interactivity, and embodiment offered by VR-based pain management is therefore distinct from and advantageous to common forms of distraction (i.e., bubbles; books; toys), passively watching television or movies, and playing a two-dimensional handheld video game or game console. Second, because the entire distraction process takes place within a safe, controlled, automated virtual environment^39^, VR-based pain management can be safely implemented in home settings. Third, previous researchers have developed VR pain distraction with burn patients in mind (e.g. snow world for burn dressing pain management) and preliminary positive results from pediatric and adult patient populations were published. Prior-generation computer-based VR pain distractions required large equipment costs as well as equipment setup and cleaning that required professional training, posing significant obstacles for the VR to be widely adopted in home settings.^40^ However, thanks to recent advancement in VR technology, VR-based pain distraction has evolved from expensive and cumbersome pieces of equipment to affordable, lightweight, mobile devices with sizes comparable to a smartphone.

This reduction in the size and cost of smartphone-based VR games, coupled with significantly improved system stability and, importantly, accessibility,^41^ has opened the door to using VR widely for burn dressing pain management in home settings.

Recent meta-analysis and reviews of published studies in the past three decades have provided evidence that VR can effectively distract patients to reduce pain and anxiety across many settings.^33,37,42-44^ However, prior studies have not investigated the feasibility and barriers of VR games for pain management during at-home burn dressing changes. Furthermore, almost all of the existing studies used computer-based VR that is technologically and financially inaccessible to patient families for everyday use in the home.

In order the address gaps in previous research, our study aimed to 1) Examine the effect of VR pain alleviation tool (VR-PAT) on reducing pediatric burn patients’ perceived pain during at-home dressing changes, 2) Examine the effect of VR-PAT on reducing pediatric patients’ perceived pain during repeated home burn dressing changes, and 3) Examine the usability of VR-PAT during pediatric dressing changes in a home setting.

## Methods

This randomized control trial tested whether a smartphone VR-PAT effectively reduced pain during repeated pediatric burn dressing changes at home. We also qualitatively assessed the ease of setup and enjoyment of the program. From September 1, 2019 to May 30, 2021 (the end of the funding period), 35 patients were recruited from the outpatient burn clinic at Nationwide Children’s Hospital (NCH) and randomly assigned to either the VR group or the control group which used standard home distraction techniques. Inclusion criteria were (1) pediatric burn patients (5-17 years) who were receiving their first outpatient dressing change at the Nationwide Children’s Outpatient Burn Clinic, (2) have a dressing that requires daily changes at home for at least one week after their first NCH outpatient clinic dressing change, and (3) can communicate orally. Exclusion criteria include: (1) Any wounds that may interfere with study procedures, (2) Vision, hearing, or cognitive/motor impairments preventing valid administration of study measures, (3) History of motion sickness, seizure disorder, dizziness, or migraine headaches precipitated by visual auras, (4) Minors in foster care (5) Suspected child abuse, (6) Unable to communicate in English, or (7) Families who do not have access to a smartphone (due to the VR-PAT game requirements).

Over the study period, 313 patients were screened, and we found 145 patients to be eligible for all factors before knowing the dressing change and 65 of these patients meeting our criteria after learning the dressing type. A trained researcher approached 49 patients for participation and 35 patients consented/assented to this RCT. Of those recruited, 24 participants returned their surveys and completed the study. No other subjects were excluded from the study following recruitment (Figure 1).

**Figure 1:**
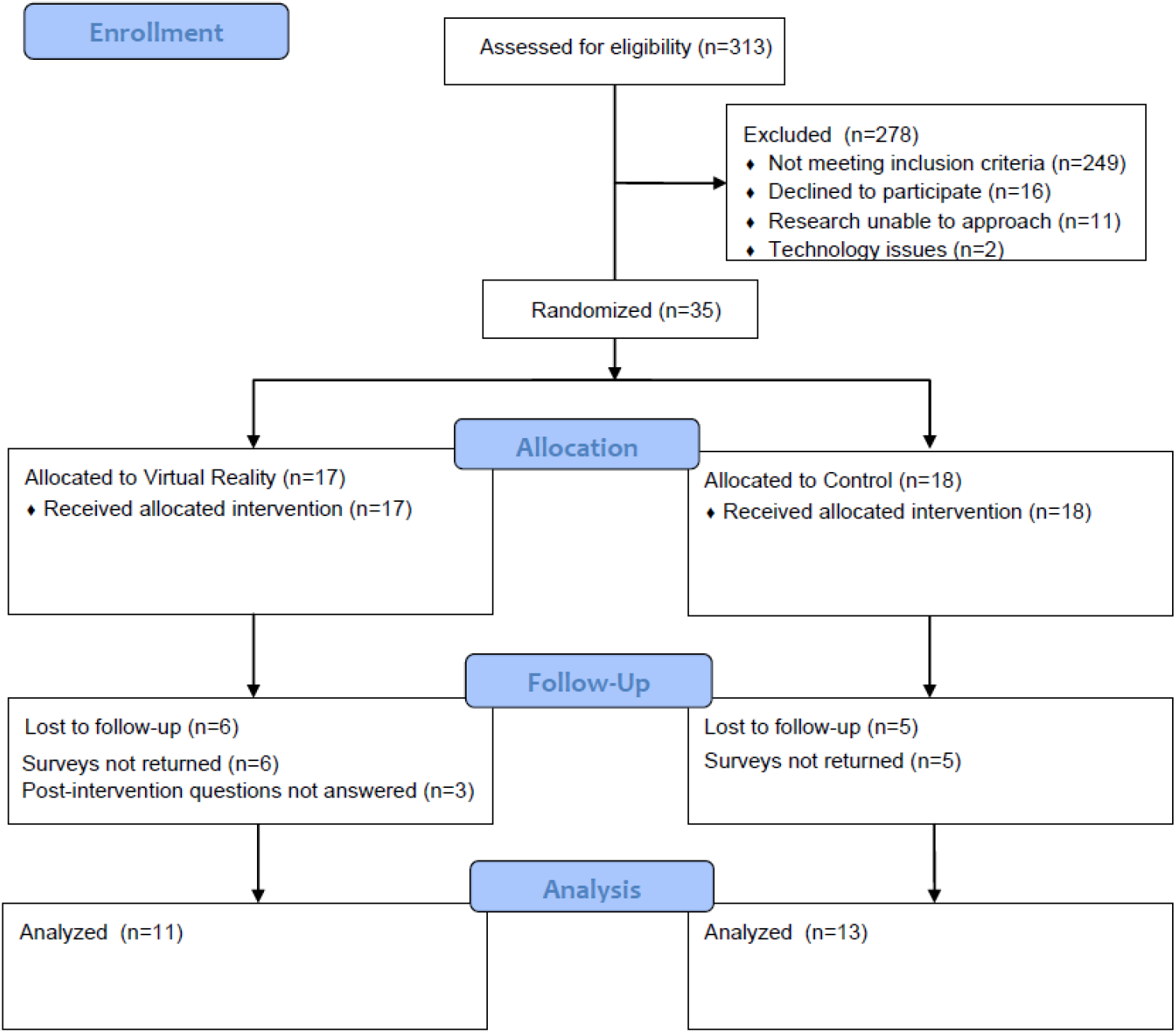
CONSORT Flow Diagram

### Study Procedures

Potential participants were identified via the Electronic Medical Record System and approached by a trained researcher at NCH. After obtaining informed consent (and assent for participants age ≥9 years), participants were asked baseline questions about their experience playing video games (days per week playing VR, console, or computer games) before being randomly assigned to either the VR-PAT or a standard of care control group using a 1:1 randomization scheme in Research Electronic Data Capture (REDCap) designed for our project.^45,46^ Every participant in the study was given a VR headset to bring home and the control group was instructed to complete the first week of dressing changes without the VR device. Participants and guardians were given up to eight surveys, to be completed each time a dressing change was necessary for up to one week and these surveys were then returned by mail in a pre-paid, self-addressed envelope

### Child Surveys (self-reported)

Participants self-rated overall pain, worst pain, and time spent thinking about pain on a scale of 1-10 (higher score means more pain). Patients in the VR group were also asked to rate their happiness, fun, engagement, and realism of the game on a scale of 1-10 (higher score means more helpful). They were also asked to report if the game made them feel not well.

### Guardian Surveys (self-reported)

Guardians were also asked to report the participants overall pain and worst pain on a scale of 1-10 (higher score means more pain). Those in the VR group were asked to report time spent using the VR-PAT, whether the participant declined to use the VR, number of voluntary interruptions, whether the device was helpful and easy to use, and any pain medications used for the burn.

Finally, participants in the VR group were contacted after a week of home dressings to ask post-intervention questions about what they liked about the game, didn’t like about the game, and whether any part of the game or set-up was too hard.

Demographic information was pulled from the electronic medical record. This included date of birth, gender, race, ethnicity, burn date, visit date, percent total body surface area (TBSA), burn severity (1^st^, 2^nd^, or 3^rd^ degree), and body area burned.

### Interventions

#### VR-PAT group

Our VR-PAT consisted of a lightweight VR headset with a Virtual River Cruise game that is played on a smartphone. In our pilot study,^47^ we found that active VR (interacting with VR game) was significantly more beneficial than passive VR (watching the same VR game). Due to these findings, only active VR was used for our intervention group for this study. VR-PAT is a standalone game developed by the Research Information Solutions and Innovation department at NCH and could be downloaded onto participants’ smartphones using either a QR code or a dedicated website.

The Virtual River Cruise game involved piloting a boat towards snow-emitting statues on a riverbank. Additionally, a thermometer on the boat would drop in temperature to create a cooling effect whenever statues were collected, and a scoreboard next to the thermometer showed children the number of statues they had hit to provide reinforcement. Children steered the virtual boat by tilting their heads, creating an immersive and active experience without interfering with the burn dressing process.

#### Control Group

Our standard of care group was able to use any distraction available in the home, including toys, video games, mobile phone, and books. The control group asked not to use the VR device during the first week of dressing changes but encouraged to use it for any dressing following the study week.

### Study Outcomes and Confounding Variables

#### Primary Outcome

Our primary outcome was pain associated with burn dressing changes. Pain scores were compared to subsequent surveys over the following week during repeated dressing changes. Secondary outcomes were time spent thinking about pain and caregiver-reported pain, both rated on a scale of 1-10. We also provided an opportunity for user feedback on VR’s effectiveness and areas for improvement.

#### Exploratory Outcome

Both children and caretakers were asked to describe the perceived enjoyability and potential adverse effects of the VR. Additionally, patients were asked questions about prior experience with VR and other gaming systems.

### Statistical Analysis

Demographic and burn characteristics were described using frequencies and percentages for the categorical variables and means and standard deviation (SD) for continuous variables. Mean, SD, and median were calculated for the primary outcome of reported pain (worst pain, overall pain, and time spent thinking of pain) across dressing changes. Child satisfaction (realism, engagement, happiness, and fun) was calculated as a mean across dressing changes. Qualitative data was collected at follow-up on the child’s utilization experience and are reported as frequencies and percentages for how often these issues were reported. Data analyses were conducted in SAS version 9.4 (SAS Institute).

## Results

The majority of children in this study were male (n=19), making up 64.7% of the VR group and 44.4% of the control group (Table 1). Most participants were also White, making up 88.2% of the VR group and 77.8% of the control group. Participants in the control group were slightly older, having a mean of 12.3 years compared to 10.7 years in the VR group. Both groups had small burns (median 1% TBSA) and 16 participants in each group had a 2^nd^ degree burn. Participants in both groups played console and computer games before but had little to no experience with VR games.

**Table 1:** Demographics, burn characteristics and prior experience with games of study participants.

| Characteristics | Intervention Group |  |
| --- | --- | --- |
|  | VR (n=17) | Control (n=18) |
| **Demographics** |  |  |
| Gender, n (%) |  |  |
| Male | 11 (64.7) | 8 (44.4) |
| Female | 6 (35.3) | 10 (55.6) |
| Race, n (%) |  |  |
| White | 15 (88.2) | 14 (77.8) |
| Black | 2 (11.8) | 3 (16.7) |
| Other | 0 (0) | 1 (5.6) |
| Age in years, mean (SD) | 10.7 (2.9) | 12.3 (3.3) |
| **Burn characteristics** |  |  |
| Burn degree, n (%) |  |  |
| Second | 16 (94.1) | 16 (88.9) |
| Third | 1 (5.9) | 2 (11.1) |
| TBSA (%), median (IQR) | 1 (1 - 2) | 1 (0.5 - 1.5) |
| **Experience with games, median (IQR)\*** |  |  |
| VR weekly | 0 (0 - 0) | 0 (0 - 0) |
| Console weekly | 2 (0 - 5.5) | 2 (0 - 7) |
| Computer weekly | 7 (2 - 7) | 5 (2 - 7) |
| Abbreviations: TBSA, total body surface area; VR, virtual reality; n, frequency; SD, standard deviation; IQR, Inter-quartile range |  |  |
| *Days per week playing games on VR, Console (i.e. PlayStation®, Xbox, Nintendo Switch™), or Computer (including mobile platforms) |  |  |

Of the 24 subjects who returned surveys, 11 were in the VR group and 13 were in the control group. There were 2 subjects who did not return medication surveys (Table 2). More subjects in the VR group reported using pain medications for the burn injury in dressings 1-5 than subjects in the control group but did not use any medications after dressing number 5. Of all the medications used, the vast majority were over the counter medications such as acetaminophen or ibuprofen.

**Table 2:**
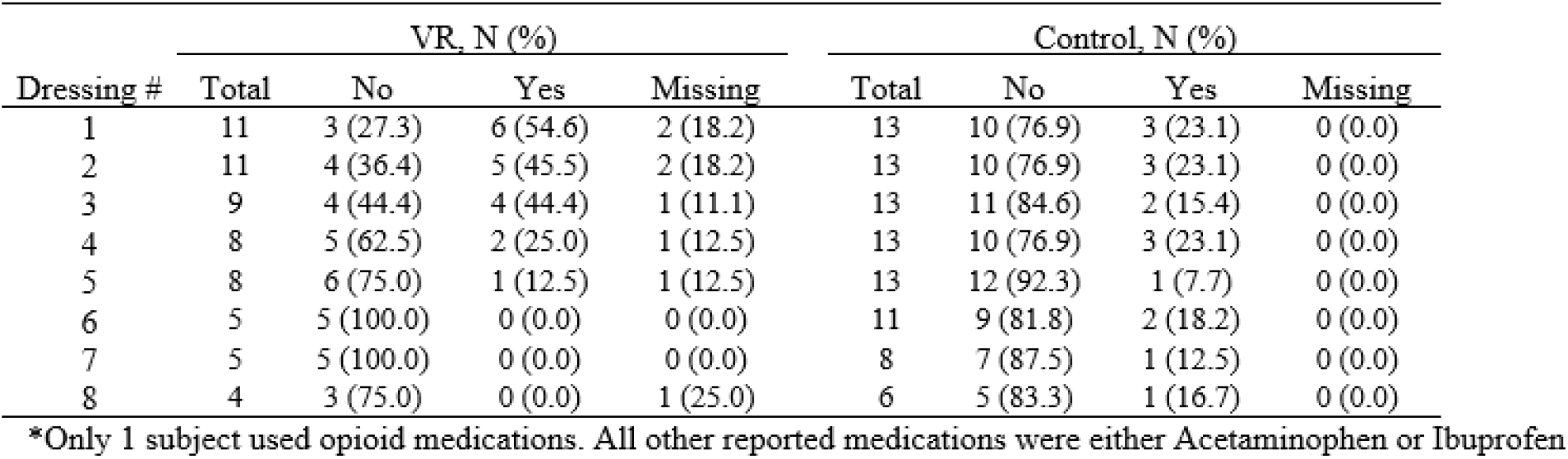
Reported pain medication use by dressing number and intervention group.

| Dressing # | VR, N (%) |  |  |  | Control, N (%) |  |  |  |
| --- | --- | --- | --- | --- | --- | --- | --- | --- |
|  | Total | No | Yes | Missing | Total | No | Yes | Missing |
| 1 | 11 | 3 (27.3) | 6 (54.6) | 2 (18.2) | 13 | 10 (76.9) | 3 (23.1) | 0 (0.0) |
| 2 | 11 | 4 (36.4) | 5 (45.5) | 2 (18.2) | 13 | 10 (76.9) | 3 (23.1) | 0 (0.0) |
| 3 | 9 | 4 (44.4) | 4 (44.4) | 1 (11.1) | 13 | 11 (84.6) | 2 (15.4) | 0 (0.0) |
| 4 | 8 | 5 (62.5) | 2 (25.0) | 1 (12.5) | 13 | 10 (76.9) | 3 (23.1) | 0 (0.0) |
| 5 | 8 | 6 (75.0) | 1 (12.5) | 1 (12.5) | 13 | 12 (92.3) | 1 (7.7) | 0 (0.0) |
| 6 | 5 | 5 (100.0) | 0 (0.0) | 0 (0.0) | 11 | 9 (81.8) | 2 (18.2) | 0 (0.0) |
| 7 | 5 | 5 (100.0) | 0 (0.0) | 0 (0.0) | 8 | 7 (87.5) | 1 (12.5) | 0 (0.0) |
| 8 | 4 | 3 (75.0) | 0 (0.0) | 1 (25.0) | 6 | 5 (83.3) | 1 (16.7) | 0 (0.0) |

\*Only 1 subject used opioid medications. All other reported medications were either Acetaminophen or Ibuprofen

Subjects in the VR group completed at least 2 dressing changes while subjects in the control group completed at least 5 dressing changes (Table 3). In the VR group, the mean child reported worst pain ranged from 3.6 (standard deviation (SD) 2.7) at the 1st dressing to 0.3 (SD 0.5) at the 8th dressing and in the control group, the range was 3.0 (SD 2.6) at the 1st dressing to 2.3 (SD 2.7) at the 8th dressing. Overall pain ranged from mean 3.2 (SD 2.4) at the 1^st^ dressing to 0.3 (SD 0.5) at the 8^th^ dressing in the VR group and 2.9 (SD 2.3) at the 1^st^ dressing to 2.2 (SD 2.4) at the 8^th^ dressing. The mean time spent thinking about pain ranged from 3.6 (SD 4.1) at the 1^st^ dressing to 0.3 (SD 0.3) at the 8^th^ dressing in the VR group and 3.5 (SD 3.6) at the 1^st^ dressing to 1.7 (SD 2.1) at the 8^th^ dressing in the control group. Children in the VR group reported less pain following the 4^th^ dressing across worst pain, overall pain, and time spent thinking about pain (Figure 2).

**Table 3:** Child reported pain by dressing number and intervention group.

| Dressing # | VR |  |  |  | Control |  |  |  |
| --- | --- | --- | --- | --- | --- | --- | --- | --- |
|  | N Obs | Mean | Standard Deviation | Median | N Obs | Mean | Standard Deviation | Median |
| **Worst Pain** |  |  |  |  |  |  |  |  |
| 1 | 11 | 3.6 | 2.7 | 3 | 13 | 3.0 | 2.6 | 3 |
| 2 | 11 | 4.3 | 2.7 | 5 | 13 | 3.3 | 2.5 | 3 |
| 3 | 9 | 2.8 | 2.1 | 3 | 13 | 2.7 | 2.7 | 2 |
| 4 | 8 | 2.8 | 2.8 | 2 | 13 | 2.6 | 3.0 | 2 |
| 5 | 8 | 1.0 | 1.5 | 0 | 13 | 1.9 | 2.8 | 1 |
| 6 | 6 | 0.7 | 1.2 | 0 | 11 | 2.6 | 3.2 | 1 |
| 7 | 5 | 0.2 | 0.5 | 0 | 8 | 2.8 | 3.1 | 2 |
| 8 | 4 | 0.3 | 0.5 | 0 | 6 | 2.3 | 2.7 | 2 |
| **Overall Pain** |  |  |  |  |  |  |  |  |
| 1 | 11 | 3.2 | 2.4 | 7 | 13 | 2.9 | 2.3 | 6 |
| 2 | 11 | 3.0 | 2.3 | 7 | 13 | 2.7 | 1.9 | 6 |
| 3 | 9 | 2.1 | 1.7 | 5 | 13 | 2.2 | 2.5 | 9 |
| 4 | 8 | 2.4 | 2.4 | 8 | 13 | 2.1 | 2.5 | 8 |
| 5 | 8 | 0.6 | 0.8 | 2 | 13 | 1.8 | 3.1 | 10 |
| 6 | 6 | 0.5 | 0.8 | 2 | 11 | 2.1 | 2.9 | 8 |
| 7 | 5 | 0.2 | 0.5 | 1 | 8 | 2.3 | 2.4 | 5 |
| 8 | 4 | 0.3 | 0.5 | 1 | 6 | 2.2 | 2.4 | 5 |
| **Time Spent Thinking about Pain** |  |  |  |  |  |  |  |  |
| 1 | 11 | 3.6 | 4.1 | 2.5 | 13 | 3.5 | 3.6 | 2 |
| 2 | 11 | 3.6 | 3.6 | 2.5 | 13 | 2.8 | 2.8 | 1 |
| 3 | 9 | 3.1 | 3.6 | 2 | 13 | 2.7 | 3.1 | 1 |
| 4 | 8 | 2.8 | 3.3 | 1.5 | 13 | 2.4 | 4.0 | 0 |
| 5 | 8 | 0.4 | 0.8 | 0 | 13 | 1.9 | 3.0 | 1 |
| 6 | 6 | 0.5 | 0.8 | 0 | 11 | 2.2 | 2.9 | 0 |
| 7 | 5 | 1.0 | 1.7 | 0 | 8 | 2.6 | 3.2 | 1 |
| 8 | 4 | 0.3 | 0.5 | 0 | 6 | 1.7 | 2.1 | 1 |

Abbreviations: N Obs, number of observations

**Figure 2:**
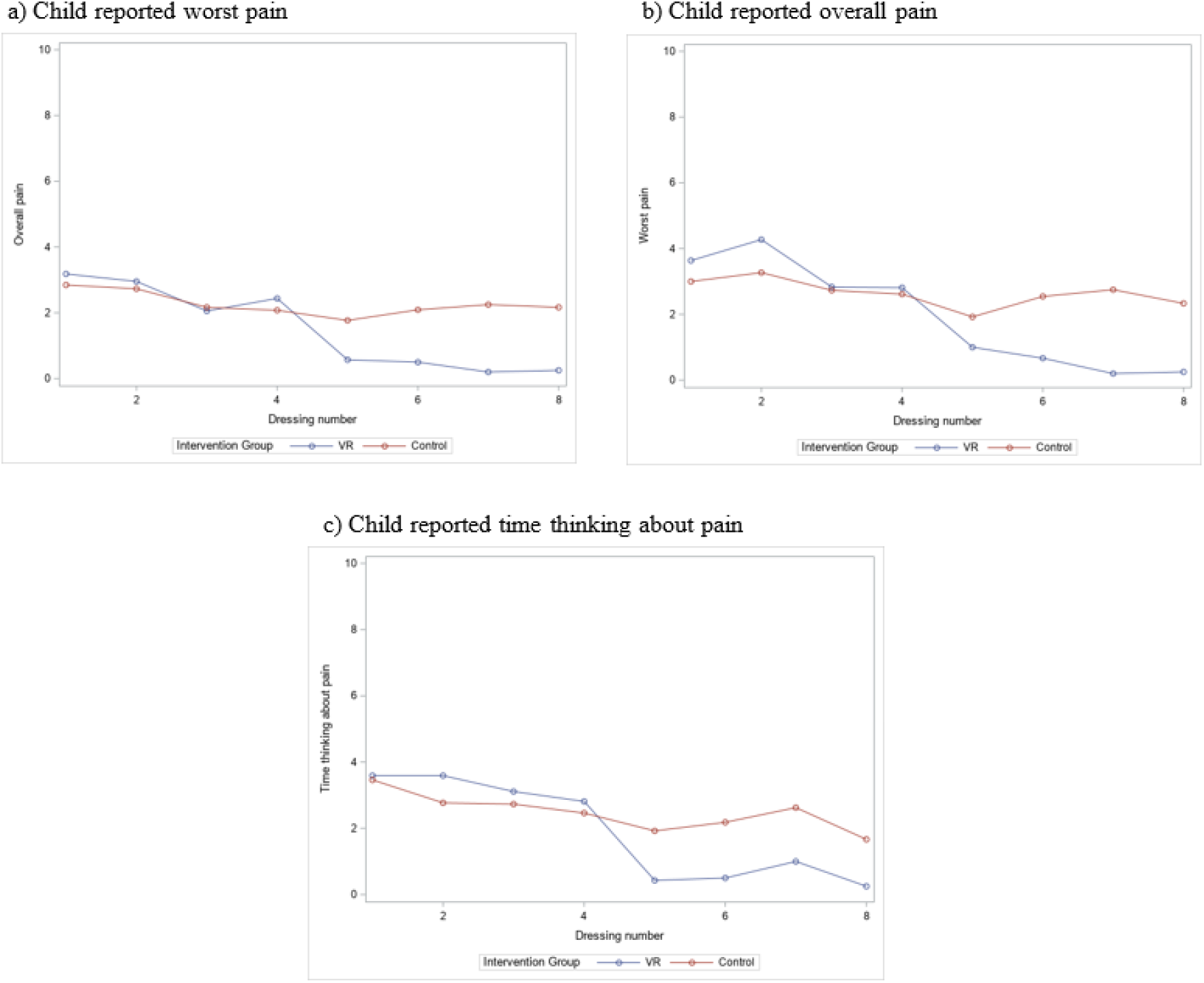
Child reported pain by dressing number and intervention group.

Caregiver reported pain followed a similar trend as the child report pain (Table 4). In the VR group, the mean caregiver reported worst pain ranged from 4.1 (SD 3.0) at the 1st dressing to 0.0 (SD 0.0) at the 8th dressing and in the control group, the range was 2.9 (SD 2.8) at the 1st dressing to 2.7 (SD 3.0) at the 8th dressing. Overall pain ranged from mean 3.2 (SD 2.6) at the 1^st^ dressing to 0.0 (SD 0.0) at the 8^th^ dressing in the VR group and 2.4 (SD 2.1) at the 1^st^ dressing to 2.3 (SD 2.6) at the 8^th^ dressing. Caregivers in the VR group also reported less pain following the 4^th^ dressing across worst pain and overall pain (Figure 3).

**Table 4:** Caregiver reported pain by dressing number and intervention group.

| Dressing # | N Obs | VR |  |  | N Obs | Control |  |  |
| --- | --- | --- | --- | --- | --- | --- | --- | --- |
|  |  | Mean | Standard Deviation | Median |  | Mean | Standard Deviation | Median |
| Worst Pain |  |  |  |  |  |  |  |  |
| 1 | 11 | 4.1 | 3.0 | 3 | 13 | 2.9 | 2.8 | 3 |
| 2 | 10 | 3.9 | 2.5 | 3 | 13 | 3.5 | 2.8 | 4 |
| 3 | 9 | 2.8 | 2.2 | 2 | 13 | 3.0 | 2.9 | 3 |
| 4 | 8 | 2.7 | 2.5 | 2.3 | 13 | 2.5 | 3.0 | 2 |
| 5 | 7 | 1.0 | 1.4 | 0 | 13 | 2.0 | 3.2 | 1 |
| 6 | 6 | 0.2 | 0.5 | 0 | 11 | 2.5 | 3.4 | 0 |
| 7 | 5 | 0.2 | 0.5 | 0 | 8 | 2.5 | 2.8 | 2 |
| 8 | 4 | 0.0 | 0.0 | 0 | 6 | 2.7 | 3.0 | 2 |
| Overall Pain |  |  |  |  |  |  |  |  |
| 1 | 11 | 3.2 | 2.6 | 2.5 | 13 | 2.4 | 2.1 | 2 |
| 2 | 10 | 3.2 | 2.1 | 2 | 13 | 2.8 | 2.2 | 3 |
| 3 | 9 | 2.3 | 1.6 | 2 | 13 | 2.2 | 2.1 | 2 |
| 4 | 8 | 2.3 | 2.0 | 1.5 | 13 | 2.0 | 2.6 | 1 |
| 5 | 7 | 0.7 | 1.1 | 0 | 13 | 1.7 | 2.9 | 0 |
| 6 | 6 | 0.5 | 0.8 | 0 | 11 | 2.0 | 2.8 | 0 |
| 7 | 5 | 0.2 | 0.5 | 0 | 8 | 2.1 | 2.3 | 2 |
| 8 | 4 | 0.0 | 0.0 | 0 | 6 | 2.3 | 2.6 | 2 |

**Figure 3:**
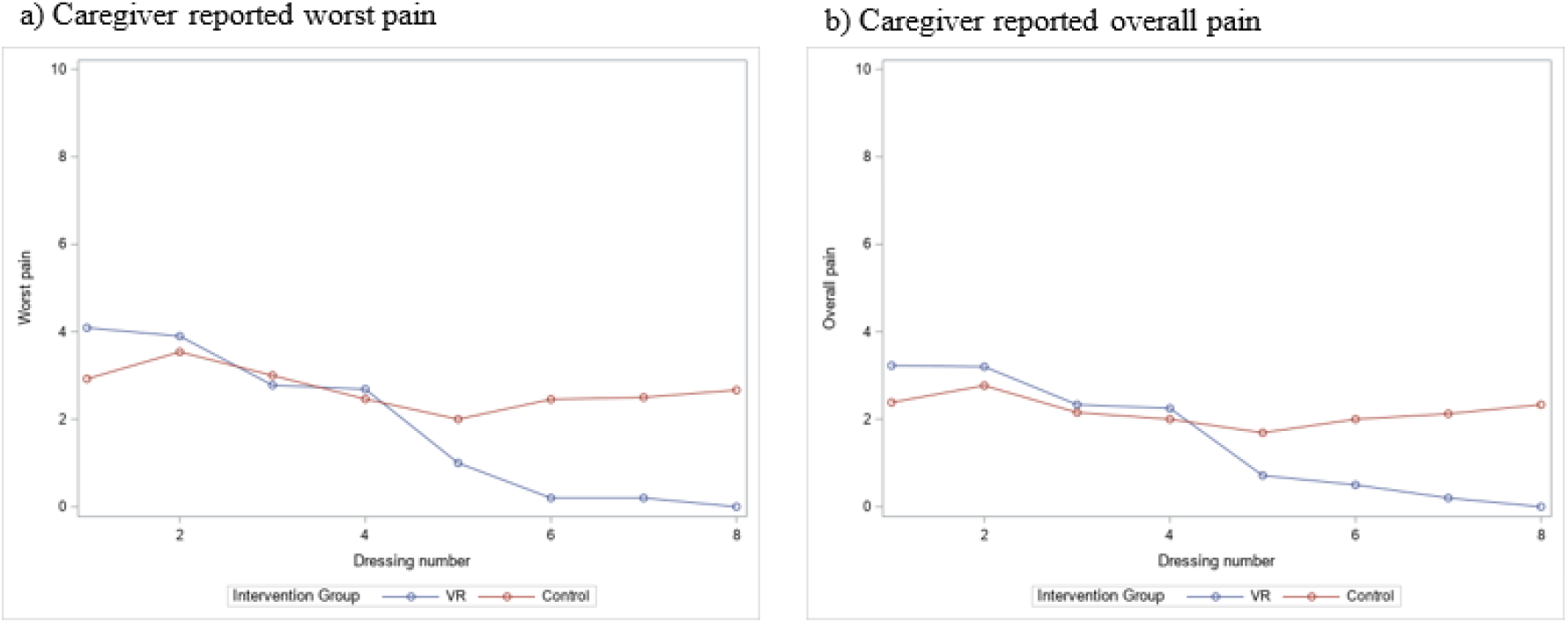
Caregiver reported pain by dressing change and intervention group.

All children in the VR group reported their satisfaction with the VR-PAT after each dressing (Figure 4). As dressing changes progressed over time, children reported increased realism (Did you feel like you were inside the game?) and engagement (How engaging did you think the game was?) with the VR-PAT. Both realism and engagement started at a mean of >5 at the first dressing and increased to >7 at the last dressing. Children’s happiness (Are you happy with the game?) and fun (How much fun did you have with it?) stayed constant at a mean of >6 across the week of dressings.

**Figure 4:**
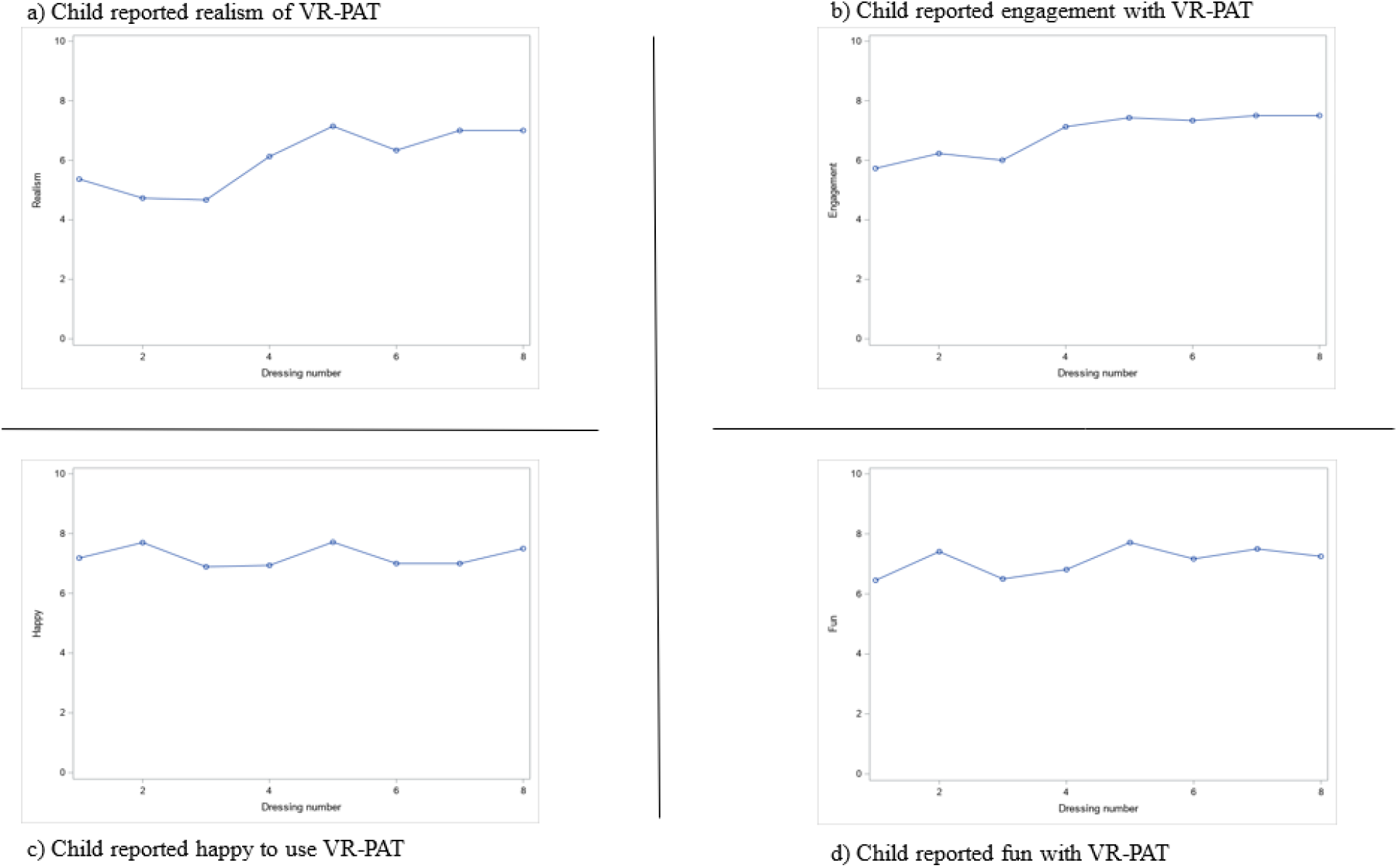
Child self-reported satisfaction with VR-PAT.

Following the week of dressing changes, children in the VR group were asked about their experience using the VR-PAT (Table 5). When asked about what they liked about the VR game, 54.5% liked the game itself, 36.4% liked that it was a distraction, and 27.3% found the VR-PAT to be calming. When asked what they did not like about the VR game, the most common responses were a desire for more levels or goals (36.4%) or nothing (27.3%). Only one child didn’t understand how to play the game at first. Finally, children were asked if there were any challenges with the VR-PAT and the majority did not express any challenges (54.5%). Of those who did report challenges, there were some technological issues with downloading the game application (18.2%), getting the system set-up (18.2%), and one found the game to be difficult to play (9.1%).

**Table 5:**
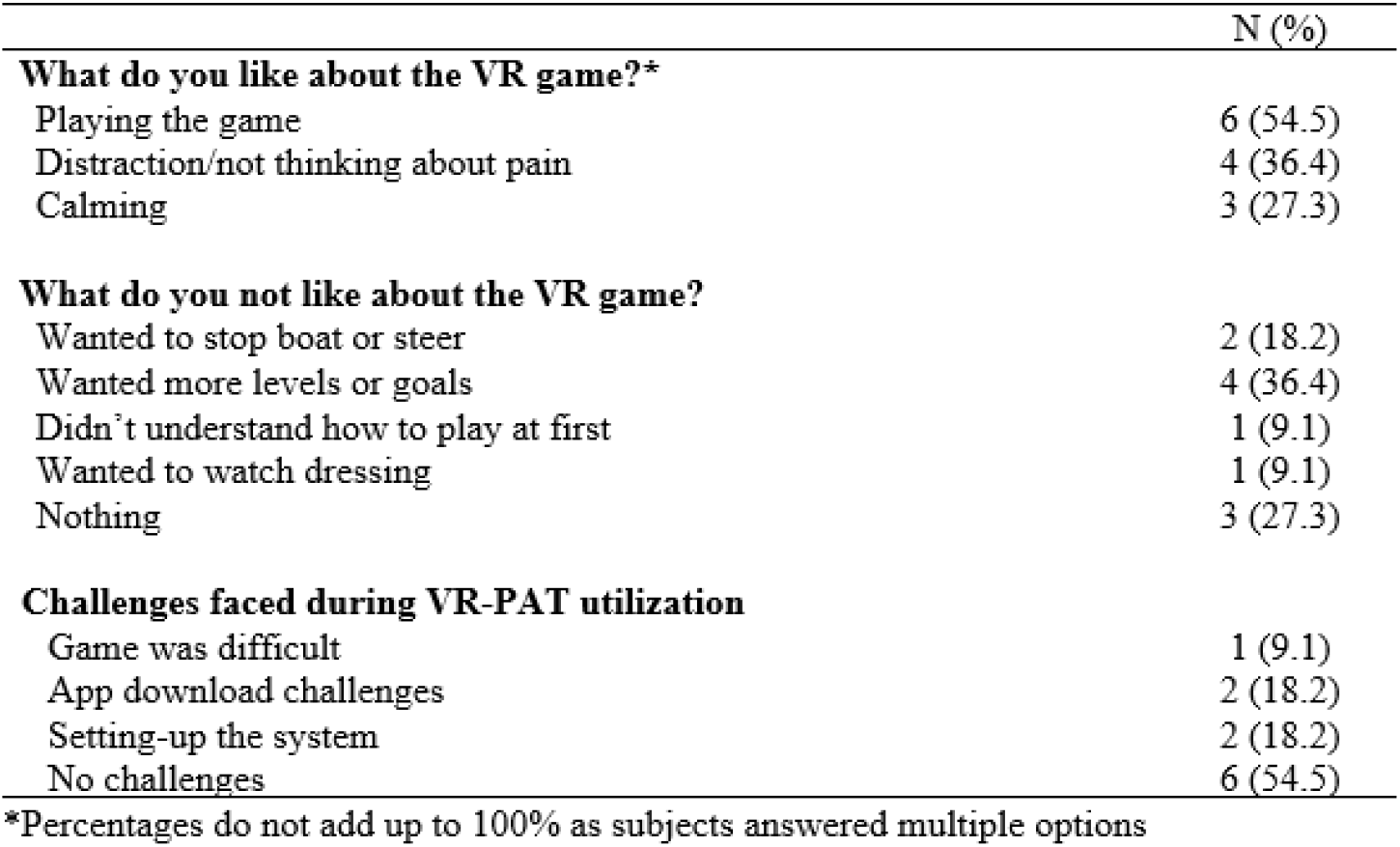
Child self-reported VR-PAT utilization experience.

## Discussion

Our study was mostly made up of male, White children with second degree burns, which is consistent with other burns studies.^1^ The number of subjects using pain medication for dressing changes was also consistent with our previous research.^47^ Interestingly, children using VR-PAT reported slightly more pain than those in the control group at the beginning of the week, but they reported less pain following the 4^th^ dressing while those in the control group stayed fairly consistent. Caregiver reported pain followed a similar trajectory across both intervention groups. All subjects did not complete the same number of surveys, so future analyses should control for dressing numbers, injury severity, age, and medication use. We also saw that children in the VR-PAT group did not report decreasing happiness or fun as the week went on and, in fact, reported increased realism and engagement. There has been some concern in the virtual reality research community that the novelty of virtual reality would wear off with increased exposure, but we found the opposite to be true in this study. This is an encouraging finding for the effectiveness of using virtual reality as a pain distraction tool for burn injuries, as these typically require more than one painful procedure. Finally, children provided valuable feedback about the usefulness of using VR at home. Subjects enjoyed playing the game and felt that it helped to be distracted from the dressing change. The things subjects did not like are important to know when either designing a VR game or choosing an existing game for this purpose. We chose to design a game that could be easily used across the age spectrum, but we learned that it may have been too simplistic, particularly for older children. Some children also prefer to be involved in the dressing change process, so having an immersive distraction is actually not preferrable to these children (one child reported this desire in our study). Most subjects in our study found the VR-PAT easy to use, but several important challenges were mentioned, particularly related to the technology and downloading of the app. The one person who found the game to be too difficult was one of the youngest participants in our study (5 years old), further justifying the lower bound of our age range for inclusion. Importantly, no children found the VR-PAT too difficult to use and stopped using because of this.

We faced several unexpected challenges during this randomized controlled trial. First, summers are the time of year where hospitals usually see the highest numbers of burn injuries in our specified age range. We missed the Summer 2019 due to difficulties in setting up the platform that would allow subjects to download the game app, which wasn’t ready until late August 2019. Second, we encountered multiple changes in Apple’s operating platform security surrounding downloading third party apps, which required our RISI team to change how the VR app could be downloaded onto an iPhone. These changes required complicated workarounds that meant we could not recruit iPhone users for periods of time. A consideration for future studies would be to host the app on Apple’s App Store or Android’s Google Play Store which makes downloading apps easier and could circumvent some of these issues. Third, we missed five months (March – August) in 2020, which included the summer, due to an NCH pause on in-person recruitment because of COVID-19. Fourth, COVID-19 resulted in institutional changes to in-person research which shifted our study to limit as much patient contact as possible. The best way to do this was to ask participants to mail their surveys back in a pre-paid, self-addressed envelope and we made three reminder calls to families. Unfortunately, we experienced a higher rate of loss to follow-up (n=10) after making this change and we attribute it to families forgetting to mail surveys and United States Postal Service (USPS) slowdowns. We believe that future studies should request surveys to be returned in person or allow families to e-mail their survey responses. Finally, there were a number of patients receiving either a long-term dressing or no dressing than we expected, which reduced the number of eligible patients. Our outpatient clinic did not have data on this prior to conducting this study, so this is something we have learned to take into consideration for future studies.

Our recommendation based on our experience conducting this study is that VR should be considered as a distraction method for home burn dressing changes. Since this is such an important topic in terms of pediatric burn care, our plan is to continue recruitment of this study until we have results for 60 subjects (n=30 per intervention group), so that we can strengthen our results and achieve the study power. We recommend that future virtual reality studies consider technology issues (like changes in smartphone operating systems), reducing loss to follow-up, and time required to recruit subjects.

## Supporting information

Author COI Forms

## Data Availability

All data produced in the present study are available upon reasonable request to the authors

## Acknowledgements

Special thanks to Rohali Keesari, PharmD, MPH who assisted with data analysis, Simon Lin MD, MBA for support with the VR game, and the nurses and staff members at the Nationwide Children’s Hospital Outpatient Burn Clinic for providing support during the patient recruitment. Without their support and help, this research study could not have been successful.

